# Individual Participant Data Network Meta-analysis of psychosocial interventions for survivors of intimate partner violence: Study protocol

**DOI:** 10.1101/2024.06.25.24309424

**Authors:** Christina Palantza, Karen Morgan, Nicky J. Welton, Hannah M. Micklitz, Lasse B. Sander, Gene Feder

## Abstract

Many systematic reviews and meta-analyses have been conducted in the field of Intimate Partner Violence (IPV) and the evidence shows small to moderate effect sizes in improving mental health. However, there is considerable heterogeneity due to great variation in participants, interventions and contexts. It is therefore important to establish which participant and intervention characteristics affect the different psychosocial outcomes in different contexts. Individual Participant Network Meta-analysis (IPDNMA) is a gold-standard method to estimate the effects with the highest precision possible and estimate moderating effects, compare the effectiveness of the different interventions and thus answer the question of which intervention is best-suited for whom. We will conduct an IPDNMA of randomised controlled trials (RCTs) of psychosocial interventions for IPV survivors aimed at improving mental health, well-being, risk-lowering and intervention acceptability outcomes compared to any type of control (PROSPERO registration number: CRD42023488502). We aim to establish a collaboration with the authors of the eligible RCT, to obtain and to harmonise the Individual Participant Data of the trials, and conduct an one-stage IPDNMA will be conducted under a Bayesian framework using the multinma package in R, after testing which characteristics of the participants and interventions are effect modifiers. There are however inherent limitations of IPDMAs, such as depending on data availability and missing nuancing through the harmonisation of variables. We aim to address these possibly by creating pseudo-IPD and sensitivity analyses. This approach is novel in the field and it can inform more efficient clinical and policy-related decision making.

## Introduction

The term Intimate Partner Violence (IPV) covers all forms of aggression towards a current or former intimate partner, including psychological, physical, sexual, or coercive controlling behaviours (1). One of the most severely impacted aspects is the mental health of the survivors (2); in fact mental disorders are more common among both men and women involved in IPV than in the general population (3). The course of IPV is intertwined with the course of mental distress (4). IPV is a worldwide public health issue (5), and it has highly adverse outcomes for victims/survivors, perpetrators and children (6,7,8). IPV was further exacerbated during the COVID-19 pandemic (9).

This increase has been consistently associated with a generally well-established risk factor for IPV, which is financial strain (10,11). Therefore, it is likely to continue rising during the current cost of living crisis, which is affecting many regions around the world (12), and non-governmental organisations working with survivors are already reporting the negative impact of the crisis (13). There is thus a heightened necessity for efficient services that are targeted to meet individuals’ needs by utilising only the resources that are needed.

Despite the heightened need, psychosocial care for survivors appears inadequate both in terms of accessibility and quality (14). The psychosocial interventions tested in research settings show some potential (15), but effect sizes are rather small (i.e. Standardised Mean Difference of −0.15 to −0.26) for major psychological outcomes such as depression, or inconsistent across reviews for safety and re-victimisation, and both clinical and statistical heterogeneity of the intervention effects are high (15,16). This could be attributed to heterogeneous populations (17). Moreover, the quality of the evidence is questionable, as many studies do not use optimal design (18), and those that do have not reported all necessary information sufficiently (16). In addition, many important aspects, such as long-term effects, harms, and differential effects for survivors with different characteristics from different contexts remain unanswered, despite the indications towards considerable impact of these differences (16,18). This is particularly problematic, because the survivors are indeed quite diverse and come from different contexts (e.g. staying with the perpetrator, staying in a shelter, co-parenting with the perpetrator) indicating different needs from psychosocial support (19).

Micklitz and colleagues categorised the tested psychosocial interventions into three main categories: psychological, advocacy-oriented and integrated (a combination of the two). Psychological interventions are psychotherapeutic interventions that employ cognitive, behavioural or emotion-focused techniques, and are usually delivered by a therapist through talking. Advocacy interventions could be summarised as advice on practical matters and continued support and counselling, enhanced access to other resources such as shelters, and facilitated safety planning (17). A realist review of advocacy interventions highlighted the critical impact of the nuances both among survivors and contexts, and the importance of matching the intervention to the survivor’s context and needs (17).

However, the hypotheses were not tested statistically, and there was considerable variation among interventions, that was not examined quantitatively. The heterogeneity of interventions has not allowed conclusions on the effectiveness to be drawn for large groups of survivors, such as mothers (20) and also blurs the evidence on certain types of interventions (21). This might also explain the contradictory evidence on effectiveness of web-based interventions (22,23). There are indications that all types of interventions have effects up to an extent for some of the possible outcomes (24), but these effects should be investigated in further detail.

A methodological approach that is well-suited for investigating such nuances in terms of population and intervention, is Individual Participant Data meta-analysis (IPDMA). This form of meta-analysis pools individual level data instead of the aggregate data from the existing studies, by requesting the datasets with the raw data from each eligible study and combining them all into one harmonised dataset (25). Meta-analyses of aggregate data are limited to the outcomes reported in publications and rely on study-level moderators lacking statistical power and are vulnerable to ecological bias (26). IPDMA overcomes these limitations by allowing outcomes and effect modifiers to be comparable across studies, enabling testing and adjusting for various within and between study moderators, both participant and intervention-related, to establish the effect sizes for different interventions and survivors (26,27,28). Importantly IPDMA has increased statistical power compared to the original studies by pooling effects. This way, the findings of (29) that participant and counselling characteristics have differential impact will be further elucidated.

Given the limited resources of health systems (30) and the different interventions tested for IPV (16), it is promising to compare the efficacy of different interventions, and identify the best-suited intervention for an individual based on their characteristics and context. A method that compares multiple different interventions and ranks their efficacy is Network Meta-analysis (NMA) (31). This is achieved by forming a network of all the interventions that have been tested for an outcome and integrating direct and indirect evidence from direct comparisons of interventions (i.e. compared in the same trial) with indirect ones (i.e. two interventions have not been tested in the same trial, but they were tested against the same control condition), thus allowing for estimation of relative effects of pairs of interventions and hierarchical ranking (32). The key assumption is that all included studies do not differ in terms of factors that could impact the relative effects (32). An issue can be inconsistency/imprecision, which occurs when the direct and indirect evidence give contradictory results, and the use of IPD can help with increasing precision and validity of the estimates (33).

Individual Patient Data Network Meta-analysis (IPDNMA), which combines IPDMA to explore individual and study-level moderators, and NMA to compare multiple interventions, provides the tools to compare different interventions for different populations (34).

### Aim and research questions

We will conduct an IPDNMA, an aggregate data network meta-analysis (NMA), a conventional meta-analysis, and an IPDMA of the overall efficacy of psychosocial interventions, to provide reliable and robust estimates of interventions and to examine which psychosocial intervention suits individual IPV survivors best to improve psychosocial and safety outcomes. More specifically the following research questions will be answered:

1. What is the overall efficacy and acceptability of psychosocial interventions in mental health and safety of IPV victims/survivors (conventional meta-analysis)?
2. What is the precise overall efficacy and acceptability of psychosocial interventions in achieving clinically relevant improvement or deterioration in the mental health and safety of IPV victims/survivors (IPDMA)?
3. Which psychosocial intervention is comparatively the most effective and acceptable in improving mental health and safety of IPV victims/survivors (NMA)?
4. Which victim/survivor and intervention characteristics moderate the efficacy and acceptability of psychosocial interventions in improving mental health and safety of IPV victims/survivors (IPDMA)?
5. Which victim/survivor and intervention characteristics moderate the comparative effectiveness and acceptability of each different psychosocial intervention in improving mental health and safety of IPV victims/survivors (IPDNMA)?

## Materials and Methods

### Design

This study aims to provide more precise and less biased estimates of the clinically relevant benefits and harms, as well as moderators to that, of each type of psychosocial intervention comparatively by obtaining the IPD of the studies to enable exploration and adjustment of effect modifiers. For these purposes, an IPDNMA will be conducted, and also a conventional meta-analysis, an aggregate data network meta-analysis, and an individual participant data meta-analysis, to compare the results and investigate the differences. The study is an update and extension of the systematic review of Micklitz and colleagues.

### Protocol registration

The study protocol has been preregistered on PROSPERO (CRD42023488502) and a project has been created on the Open Science Framework https://osf.io/72uwe/?view_only=1ba290a378514c3a929d7eac035bfd67. The Preferred Reporting Items for systematic reviews and meta-analyses (PRISMA) statements for systematic review protocols (PRISMA-P) (35) are followed (see appendix).

### Eligibility criteria

The eligibility criteria based on the PICO (Participants, Interventions, Comparators and Outcomes) are listed in Table 1.

**Table 1.** Eligibility criteria by PICO.

|  |  |
| --- | --- |
| **Participants** | Individuals or couples reporting either past or ongoing IPV at randomisation, but studies solely on those involved in perpetration are excluded |
| **Interventions** | Psychosocial interventions aimed at IPV survivors using any mode of delivery at any setting. Both psychotherapeutic and advocacy interventions are deemed as psychosocial. Interventions aimed solely at perpetrators, primary prevention to individuals who have not experienced IPV at baseline, non-psychological risk factors such as sexual behaviour or financial situation, and physical safety (such as being in a refuge/shelter and police interventions) are not included. |
| **Comparators** | All comparators are eligible, including no intervention, waiting list, Treatment as Usual (TAU), another active intervention, placebo or sham version of the intervention. The interventions will be categorised and classified into nodes of the network based on their description regardless of whether they were tested as control or active condition in the original studies. |
| **Outcomes** | Eligible outcomes are all outcomes related to mental health and well-being (e.g. depression, quality of life). |

Only randomised controlled trials (RCTs) are eligible for inclusion, including pilot RCTs and studies with few participants, as they will be merged with larger studies testing a similar intervention. All papers published in peer-reviewed journals are eligible, as well as protocols and trial registrations in case the authors already have data that they can provide. No language or publication date restrictions are applied.

### Identification of studies-information sources

A search strategy with keywords and phrases such as “intimate partner violence” and “psychotherapy” will be used to search the Web of Science database from inception to late November 2023 (see appendix for full search string). This search strategy was used by Micklitz and colleagues, so an update of their search will be performed in the databases they searched (PsycINFO, Medline, Embase, and Cochrane Central). Clinical trial registries will be searched, and forward and backward search will be performed.

### Study selection process

Two independent researchers will screen the titles and abstracts. All completed studies included in the systematic review of Micklitz et al will be included. The authors of all the protocols and trial registrations identified by Micklitz and colleagues will be contacted to find out whether data are available, and if they agree to share their data, they will be offered co-authorship of the publications derived from the study. If the studies or protocols included at this stage have been published, the full text will be screened by the two independent researchers. In the case of trial registrations, the complete registrations will be screened by the two independent researchers, or a detailed description will be requested from the principal investigators of the studies. In case of disagreement between the two researchers, a third, senior member of the team will be consulted. The screening and inclusion of abstracts and full texts will be done using Rayyan (36).

### Ethical considerations

Since this type of study is essentially a secondary analysis of existing data, it does not need to be ethically approved. However, the trialists of the eligible RCTs are responsible for the ethical re-use of the data they collected, so it is up to them to either consult their local Ethical Review Boards and possibly to submit this re-use for ethical approval, or to request us to sign a Data Sharing Agreement with them. The secure handling of this sensitive type of data is described in the following section “Data Management”.

### Data management

A collaborative project will be created on the Research Data Storage Facility of the University of Bristol, managed by GF and accessible to the rest of the authors. The data can be uploaded to that space by the authors of the primary studies. This cloud is compliant with the General Data Protection Regulation. In case the authors face unresolvable issues with this cloud, secure sharing links to a private folder at the institutional OneDrive of CP will be generated. As soon as the data have been uploaded, they will be saved directly in the RDSF and removed from OneDrive. The data will be copied to the university-managed and password protected device of CP for analysis. Since the data are sensitive, a Data Protection Impact Assessment will be performed.

### Data extraction of study characteristics and aggregate data

The study characteristics to be extracted in a spreadsheet are: author, publication year, citation, country, recruitment setting, trial setting, number of trial arms, number of follow-ups, timing of follow-ups, timing of post-treatment, primary and secondary outcomes, type of randomisation, unit of allocation, method of randomisation, sample size in total and in each arm, treatment and study dropout, intention-to-treat or per-protocol analysis, age, gender, current frequency and/or severity of IPV, past frequency and/or severity of IPV, specific target population, eligibility criteria, description of intervention, theoretical orientation, delivery mode, type of therapists/facilitators, number and duration of sessions, frequency of sessions, description of control. The characteristics and aggregate data extracted by Micklitz et al (16) will be used.

### Data collection

The corresponding authors of eligible studies will be emailed and invited to share the individual-level data of their study. The most recent email address of the author will be sought. If necessary, a reminder will be sent after three weeks and six weeks. If unreachable, the same process will be followed for the senior author of the study. If no valid email address of neither the corresponding nor the senior author can be found, social networking websites such as ResearchGate and LinkedIn will be used to reach the authors, and the same number of reminders will be sent if necessary. The IPD will be deemed unavailable if no response is received after three weeks of the second reminder to the senior author (12 weeks in total). After authors have accepted to share the IPD, there is no specific time limit to send the data.

### Data items

It is expected that not all eligible studies have collected data on all possible moderating characteristics, such as income level, and in such a multifaceted issue, there can be various potentially impactful variables. Therefore, the complete datasets will be requested including baseline and all available follow-ups, with an exception for variables that could lead to the identification of a participant and violate anonymity.

### Data harmonisation

A standard coding of the characteristics of participants will be established based on the operationalisation used by the majority of the studies and the data of each individual study will be recoded accordingly. We currently propose the operationalisation below, but the final one will depend on the data. If a study has not collected any data on some of the listed demographic or socio-economic characteristics, this will be taken into account in the quality assessment.

1. Biological sex: female/male/intersex
2. Gender identity: cisgender/transgender/gender non-conforming
3. Sexual orientation: heterosexual/homosexual/bisexual/other
4. Age at baseline: continuous variable
5. Relationship status: being in a relationship/not being in a relationship/not sure
6. Cohabitation status: cohabiting with partner/non-cohabiting with partner
7. Parenthood: parent living with children/parent having some contact with their children/ parent having no contact with their children/non-parent
8. Education: Uneducated / Primary education/ Secondary education/ Tertiary education/Other
9. Employment status: student/employed/unemployed seeking employment/unemployed not seeking/retired or disability
10. Income level: OECD (Organisation for Economic Co-operation and Development) quintiles
11. Ethnicity: categorical variable with the respective ethnicities in the respective country/trial
12. Presence of psychiatric/developmental disorder
13. Presence of history of abuse in the family
14. Presence of substance dependence

The data from different validated scales will be divided by the standard deviation. If a study uses more than one scale for the same outcome, these scales will be combined in a sort of “within-study-synthesis”. If any scale on characteristics of the therapists, such as level of empathy, is available, it will be used.

### IPD integrity

In order to check the integrity of the data received, a replication of the summary statistics and main analysis of each study will be attempted by two researchers, and if it fails, the authors will be asked for clarification. In addition, conventional MA with the aggregate data will be performed to replicate the existing findings, and the statistical package that we will use (multinma) accounts for differences by IPD availability status. More details are provided in the statistical analysis section.

### Risk of bias and quality assessment

The risk of bias of the studies will be assessed using the Cochrane Risk of Bias Assessment 2.0 (37), which assesses bias arising from the randomization process, bias due to deviations from the intended interventions, bias due to missingness of outcome data, bias in the measurement of outcome, and bias due to selective reporting of outcomes. Publication or small sample bias will be examined through visual inspection of the funnel plots for each outcome. The overall quality of the evidence will be assessed with the Grading of Recommendations Assessment, Development and Evaluation (GRADE)—Guidelines (38), which, apart from the risk of bias, assesses inconsistency, indirectness, imprecision and overall quality of the evidence. These assessments will be performed by two independent researchers and disagreements will be resolved through discussion with a third more senior researcher. For the studies already assessed by Micklitz and colleagues (16), the domains of bias arising from missing data and selective outcome reporting will be revisited in case the IPD can elucidate previously unclear points. Publication bias will be assessed through a funnel plot and Egger’s test.

### Categorisation of interventions and control conditions

Based on the work of Micklitz and colleagues, the interventions that have been tested so far can be categorised into the following nodes:

1. Advocacy interventions: interventions based on the empowerment theory of Dutton (39) and the stages of change model (40), where facilitators use non-directive techniques such as motivational interviewing to guide the survivors to reach specialised services (also legal and financial ones), and to establish a safety plan
2. Integrative interventions: interventions combining the elements of the advocacy interventions with those of the psychological ones, including safety planning
3. Cognitive behavioural interventions: psychological interventions employing cognitive and behavioural techniques, such as cognitive restructuring
4. Third wave therapies: psychological interventions with more targeted cognitive approaches, such as mindfulness
5. Systemic interventions: family or couple based psychological interventions
6. Other psychological interventions: other psychological approaches tested by a small number of studies, such as interpersonal, humanistic or mindfulness/mediation based

The control conditions are expected to be categorised for the NMA as follows, unless different clusters of control conditions with common elements appear in the final set of eligible studies:

1. Waiting list
2. No intervention
3. Information on services
4. Referral to services
5. Active psychiatric care
6. Contact
7. Community care
8. Support groups
9. Hotlines

However, small changes might be made depending on the elements included in the control conditions of the included studies.

### Outcomes and effect measures

There is a vast array of psychosocial, risk lowering and safety-behaviour outcomes of interest tested by the primary studies, which is not ideal to establish the effectiveness of interventions with certainty, but it is the reality of the field (41). Psychosocial interventions use primarily psychological techniques that target mental health symptoms (16). So, in our study, the primary outcomes are some of the most commonly measured psychological distress symptoms of

1. depression
2. anxiety
3. PTSD

The secondary psychosocial outcomes are: suicidal ideation, substance use, perceived self-efficacy, self-esteem, quality of life, social support, decisional conflict, empowerment, space for action, all as scores in validated self- or clinician-rated scales, clinical interviews, or composites of scales. The main timepoint is post-treatment, but all available follow-ups will be extracted, and if there is sufficient homogeneity in the timing of follow-ups, analyses will be conducted.

Additionally, the risk-lowering outcomes are: continuation of frequency/intensity of IPV, as a whole (using the total scores of scales), and also the subtypes of psychological, physical, sexual abuse and coercive control separately when available (as scores on subscales), which will all be operationalised continuously as scores on validated (sub)scales. Safety-related behaviours will be examined only if there is not too much heterogeneity in the definition and operationalisation of this outcome by the primary studies.

It is also interesting to investigate the effects of the interventions on service use, and acceptability of the interventions. Service use will be operationalised as scores on checklists. Satisfaction with the treatment will also be operationalised as scores on respective questionnaire items. Acceptability of the interventions will be defined as the number of participants dropping out of the intervention (not necessarily the whole study and assessments) (dichotomous outcome). The effect size for all continuous outcomes will be Hedge’s g, and for all dichotomous outcomes the odds ratios will be calculated. Like for primary outcomes, the main timepoint of interest is post-treatment, but all available follow-ups will be extracted, and if there is sufficient homogeneity in the timing of follow-ups, analyses will be conducted.

### Statistical analysis-synthesis

Firstly, a conventional meta-analysis excluding studies comparing active psychosocial interventions will be conducted to establish the overall efficacy of psychosocial interventions. Secondly, an aggregate data network meta-analysis will be conducted to compare and rank the efficacy of each psychosocial intervention; the active interventions that were tested as controls in the original studies will be classified in the node of active interventions they share the same elements and approaches with. Thirdly, an Individual Participant Data Meta-analysis will be conducted (excluding the studies that compare two active psychosocial interventions) to establish the overall precise effects and moderators of efficacy of psychosocial interventions as a whole. Given the high chance that numerous potential moderators will be available, participant or intervention related, we will conduct the analysis in two steps, as demonstrated by (42). Missing data will be multiply imputed for each study using multiple imputation. If the regression coefficients are available in the publications of studies that do not provide IPD, we will create pseudo-IPD for continuous outcomes, as described in (43).

The same procedure will be repeated including all studies for the Individual Participant Data Network meta-analysis. Additionally, in the first step we will select the variables that show a higher interaction with the different nodes, by fitting a penalised linear regression model for continuous outcomes and a penalised logistic regression for dichotomous outcomes. This will be done through the lasso technique in STATA. In this step we will also test whether survivor and setting characteristics (such as being separated or still living together with their partner, and clinical or community settings) are effect modifiers or prognostic factors, and based on the results, the following stages of the analysis will be divided into subgroups of studies that are similar in effect modifiers, so that the transitivity assumption is met. In the second step, a one-stage IPDNMA will be conducted under a Bayesian framework using the multinma package in R. The characteristics of the interventions, such as number of sessions, format and mode of delivery will be tested as effect modifiers. We will also conduct a multilevel network meta-regression (using multinma in R) to enable studies providing IPD to be combined with those that do not, as demonstrated by (44). For the outcome of acceptability (defined as dropout from the intervention) we will measure an absolute treatment effect, as many control conditions are inactive, by modelling the probability of dropout through a generalised model of binomial likelihood, as in (42). Heterogeneity will be assessed by comparing the fit of fixed and random effects models, reporting between-study variance, τ^2^, and explored through inspection of regression coefficients and sub-group analyses. Inconsistency will be assessed by fitting a model that relaxes the consistency assumption, and explored further using a node-splitting model. In all of the aforementioned models where this is possible (aggregate data NMA, IPDMA, IPDNMA), if it is still appropriate with the combination of elements/components the interventions consist of, we will explore fitting IPD component NMA models (45,46,47). A sensitivity analysis excluding studies with high risk of bias will be conducted separately for each bias domain. The rankings of the interventions and the Surface Under the Cumulative Ranking (SUCRA) scores will be calculated to summarise the results of the IPDNMA. The results of all analyses will be compared to further investigate the precision of the estimates.

## Discussion

We plan to conduct an IPDNMA to estimate precisely the effects of psychosocial interventions on the mental health of IPV survivors, to compare the interventions, and to examine potential moderating effects of the characteristics of the participants and the interventions. We intend to fulfil this aim by establishing a collaboration with the authors of the RCTs and thus obtaining the datasets of the studies and combining them.

### Strengths and limitations

This methodology is considered a gold standard in evidence synthesis (27) and it is novel in the field of IPV. The synthesis approach allows research findings to be integrated to increase precision (26), whilst thoroughly investigating factors affecting the effects of the interventions on the outcomes with advanced statistical methods. These factors will pertain to individuals, interventions and contexts, and possibly their interaction, to address heterogeneity as fully as possible. The certainty of the evidence will also be assessed under the GRADE method (38).

However, there are admittedly limitations to our approach. We will be limited by the data measured in the included studies, and statistical power is bound to vary across outcomes. In addition, harmonising the outcomes across studies that have measured them in different ways means that some nuancing might be missed, but we plan to address this by sensitivity analyses. Another common issue of IPDMAs is that the IPD are often impossible to obtain (48). We plan to deal with this issue by attempting to contact the researchers through different channels and by using pseudo-IPD (43). Efforts will be made to reduce publication bias by expanding the literature search of Micklitz and colleagues, and searching trial registries and grey literature.

### Relevance

The field of IPV is highly heterogeneous, and matching individuals and contexts with interventions is crucial (17). This has not been done yet with quantitative methods that limit personal biases and offer precise estimates of the impact of each of the many factors intertwined in the provision of psychosocial support of IPV survivors. Pinpointing impactful factors and comparing the existing interventions can contribute to more efficient decision making both for practice and policy.

## Supporting information

PRISMA statement

Search strategy

## S1 Table. Eligibility criteria by PICO

### Funding statement

Christina Palantza is in receipt of an annual stipend from the University of Bristol from 2023 to 2027. This is the only funding source of this review. The funder had no role in the development of this protocol.

### Author contributions

Christina Palantza conceptualised the main aims and methods of the review, drafted the protocol. Karen Morgan and Gene Feder oversaw the development of the protocol and provided expert advice on conceptual aspects. Nicky Welton oversaw the development of the protocol and provided expert advice on methodological and statistical aspects. Lasse Sander and Hannah Micklitz conducted the systematic review and meta-analysis that is to be updated and extended by this review, reviewed the protocol and provided the materials of their review.

### Financial competing interest

None apart from CP’s stipend.

### Non-Financial competing interest

None

### Patient and Public Involvement

A Patient and Public Involvement group of survivors of intimate partner violence from the UK was consulted and provided input on conceptual aspects.

### Transparency Declaration

Christina Palantza affirms that the manuscript is an honest, accurate, and transparent account of the study being outlined; no important aspects of the study have been omitted.

## Data Availability

It will not be possible to share the data publicly upon completion of the study, because they belong to the authors that conducted the eligible studies, and they will share them with us only for the analysis of this study.

https://osf.io/72uwe/?view_only=1ba290a378514c3a929d7eac035bfd67

## Notes

### Competing Interest Statement

The authors have declared no competing interest.

### Funding Statement

The study was funded through Christina Palantza's postgraduate research stipend from the University of Bristol

### Author Declarations

This manuscript reports the protocol for secondary research using individual participant data from randomised controlled trials that have published their findings. Ethics approval is not required for secondary research. The primary trials all had ethics approval from the relevant national or institutional bodies.

