## Supplementary material for "Individual Participant Data Network Meta-analysis of psychosocial interventions for survivors of intimate partner violence: Study protocol": Search strategy

### Appendix A

| **CENTRAL** | | | **Medline** (via PubMed) | | **PsycInfo** (via Ebsco) | | **Embase** (via Ovid) | |
| --- | --- | --- | --- | --- | --- | --- | --- | --- |
|  | #1 | MeSH descriptor:  [Intimate Partner Violence]  explode all trees | 1 | intimate partner violence  [MeSH Terms] | S1 | MA Intimate Partner Violence | 1 | partner violence.sh. |
|  | #2 | MeSH descriptor: [Spouse Abuse] explode all trees | 2 | spouse abuse [MeSH Terms] | S2 | MA Spouse Abuse | 2 | domestic violence.sh. |
|  | #3 | MeSH descriptor: [Domestic Violence] explode all trees | 3 | domestic violence [MeSH Terms] | S3 | MA Domestic Violence | 3 | gender based violence.sh. |
|  | #4 | MeSH descriptor: [Gender-Based Violence] explode all trees | 4 | gender-based violence  [MeSH Terms] | S4 | MA Gender-based violence | 4 | marital rape.sh. |
|  | #5 | MeSH descriptor: [Rape]  explode all trees | 5 | rape [MeSH Terms] | S5 | MA Rape | 5 | battered women.sh. |
|  | #6 | MeSH descriptor: [Battered Women] explode all trees | 6 | battered women [MeSH Terms] | S6 | MA Battered Women | 6 | emotional abuse.sh. |
|  | #7 | (“intimate partner violence”):ti,ab,kw | 7 | “intimate partner violence” [Title/Abstract] | S7 | SU Intimate Partner Violence | 7 | “intimate partner violence”.ti. or “intimate partner violence”.ab. |
|  | #8 | (“partner violence”):ti,ab,kw | 8 | “partner violence” [Title/Abstract] | S8 | SU Dating Violence | 8 | “partner violence”.ti. or  “partner violence”.ab. |
|  | #9 | (“interpersonal violence”):ti,ab,kw | 9 | “interpersonal violence” [Title/Abstract] | S9 | SU Domestic Violence | 9 | “interpersonal violence”.ti. or “interpersonal violence”.ab. |
|  | #10 | (“dating violence”):ti,ab,kw | 10 | “dating violence” [Title/Abstract] | S10 | SU Rape | 10 | “dating violence”.ti. or  “dating violence”.ab. |
|  | #11 | (“domestic violence”):ti,ab,kw | 11 | “domestic violence” [Title/Abstract] | S11 | SU Battered Females | 11 | “domestic violence”.ti. or  “domestic violence”.ab. |
|  | #12 | (“family violence”):ti,ab,kw | 12 | “family violence” [Title/Abstract] | S12 | TI “intimate partner violence” OR AB “intimate partner violence” | 12 | “family violence”.ti. or  “family violence”.ab. |
|  | #13 | (gender?based violence):ti,kw,ab | 13 | “gender-based violence” [Title/Abstract] | S13 | TI “partner violence” OR  “AB “partner violence” | 13 | “gender*based violence”.ti. or “gender*based violence”.ab. |
|  | #14 | (“violence in close relationships”):ti,ab,kw | 14 | “gender based violence” [Title/Abstract] | S14 | TI “interpersonal violence” OR  AB “interpersonal violence” | 14 | “violence in close relationships”.ti. or “violence in close relationships”.ab. |
|  | #15 | (“intimate partner abuse”):ti,ab,kw | 15 | “violence in close relationships” [Title/Abstract] | S15 | TI “dating violence” OR  AB “dating violence” | 15 | “intimate partner abuse”.ti. or “intimate partner abuse”.ab. |
|  | #16 | (spous* abuse):ti,ab,kw | 16 | “intimate partner abuse” [Title/Abstract] | S16 | TI “domestic violence” OR  AB “domestic violence” | 16 | “partner abuse”.ti. or  “partner abuse”.ab. |
|  | #17 | (“wife abuse”):ti,ab,kw | 17 | spous* abuse [Title/Abstract] | S17 | TI “family violence” OR  AB “family violence” | 17 | “spous* abuse”.ti. or  “spous* abuse”.ab. |
|  | #18 | (“abusive relationship”):ti,ab,kw | 18 | “abusive relationship” [Title/Abstract] | S18 | TI “gender-based violence” OR AB “gender-based violence” | 18 | “abusive relationship”.ti or  “abusive relationship”.ab. |
|  | #19 | (“abusive partners”):ti,ab,kw | 19 | “marital abuse” [Title/Abstract] | S19 | TI “gender based violence” OR  AB “gender based violence” | 19 | “marital abuse”.ti. or  “marital abuse”.ab |
|  | #20 | (“marital abuse”):ti,ab,kw | 20 | “marital rape” [Title/Abstract] | S20 | TI “violence in close relationships” OR AB “violence in close relationships” | 20 | “marital rape”.ti. or  “marital rape”.ab. |
|  | #21 | (“dating abuse”):ti,ab,kw | 21 | “dating abuse” [Title/Abstract] | S21 | TI “intimate partner abuse” OR  AB “intimate partner abuse” | 21 | “dating abuse”.ti. or  “dating abuse”.ab. |
|  | #22 | (“intimate partner aggression”):ti,ab,kw | 22 | “intimate partner aggression” [Title/Abstract] | S22 | TI “spous* abuse” OR  AB “spous* abuse” | 22 | “intimate partner aggression”.ti. or “intimate partner aggression”.ab. |
|  | #23 | (“battered women”):ti,ab,kw | 23 | “intimate terrorism” [Title/Abstract] | S23 | TI “abusive relationship” OR  AB “abusive relationship” | 23 | “intimate terrorism”.ti. or  “intimate terrorism”.ab. |
|  |  |  | 24 | “battered women” [Title/Abstract] | S24 | TI “marital abuse” OR  AB “marital abuse” | 24 | “battered females”.ti. or  “battered females”.ab. |
|  |  |  | 25 | “battered wives” [Title/Abstract] | S25 | TI “marital rape” OR  AB “marital rape” | 25 | “battered women”.ti. or  “battered women”.ab. |
|  |  |  |  |  | S26 | TI “dating abuse” OR  AB “dating abuse” | 26 | “battered wives”.ti. or  “battered wives”.ab. |
|  |  |  |  |  | S27 | TI “intimate partner aggression” OR AB “intimate partner aggression” |  |  |
|  |  |  |  |  | S28 | TI “intimate terrorism” OR  AB “intimate terrorism” |  |  |
|  |  |  |  |  | S29 | TI “battered females” OR  AB “battered females” |  |  |
|  |  |  |  |  | S30 | TI “battered women” OR  AB “battered women” |  |  |
|  |  |  |  |  | S31 | TI “battered wives” OR  AB “battered wives” |  |  |
|  | **#24** | #1 OR #2 OR #3 OR #4 OR #5 OR #6 OR #7 OR #8 OR #9 OR #10 OR #11 OR #12 OR #13 OR #14 OR #15 OR #16 OR #17 OR #18 OR #19 OR #20 OR #21 OR #22 OR #23 | **26** | 1 OR 2 OR 3 OR 4 OR 5 OR 6 OR 7 OR 8 OR 9 OR 10 OR 11 OR 12 OR 13 OR 14 OR 15 OR 16 OR 17 OR 18 OR 19 OR 20 OR 21 OR 22 OR 23 OR 24 OR 25 | **S32** | S1 OR S2 OR S3 OR S4 OR S5 OR S6 OR S7 OR S8 OR S9 OR S10 OR S11 OR S12 OR S13 OR S14 OR S15 OR S16 OR S17 OR S18 OR S19 OR S20 OR S21 OR S22 OR S23 OR S24 OR S25 OR S26 OR S27 OR S28 OR S29 OR S30 OR S31 | **27** | 1 OR 2 OR 3 OR 4 OR 5 OR 6 OR 7 OR 8 OR 9 OR 10 OR 11 OR 12 OR 13 OR 14 OR 15 OR 16 OR 17 OR 18 OR 19 OR 20 OR 21 OR 22 OR 23 OR 24 OR 25 OR 26 |
|  | #25 | MeSH descriptor: [Mental Health Services] explode all trees | 27 | Mental health services  [MeSH Terms] | S33 | MA mental health services | 28 | mental health care.sh. |
|  | #26 | MeSH descriptor: [Psychotherapy] explode all trees | 28 | Psychotherapy [MeSH Terms] | S34 | MA psychotherapy | 29 | psychotherapy.sh |
|  | #27 | MeSH descriptor: [Couples Therapy] explode all trees | 29 | Couples Therapy [MeSH Terms] | S35 | MA couples therapy | 30 | couple therapy.sh. |
|  | #28 | MeSH descriptor: [Counseling] explode all trees | 30 | Counseling [MeSH Terms] | S36 | MA counseling | 31 | counseling.sh. |
|  | #29 | MeSH descriptor: [Psychosocial Intervention] explode all trees | 31 | Psychosocial intervention  [MeSH Terms] | S37 | MA psychosocial intervention | 32 | psychosocial care.sh. |
|  | #30 | MeSH descriptor: [Social Work, Psychiatric] explode all trees | 32 | Social Work, Psychiatric  [MeSH Terms] | S38 | MA social work, psychiatric | 33 | nursing intervention.sh. |
|  | #31 | MeSH descriptor: [Psychiatric Rehabilitation] explode all trees | 33 | Psychiatric Rehabilitation  [MeSH Terms] | S39 | MA psychiatric rehabilitation | 34 | web-based intervention.sh. |
|  | #32 | MeSH descriptor: [Therapy, Computer-Assisted] explode all trees | 34 | Therapy, computer-assisted  [MeSH Terms] | S40 | MA therapy, computer assisted | 35 | telehealth.sh. |
|  | #33 | MeSH descriptor: [Telemedicine] explode all trees | 35 | Telemedicine [MeSH Terms] | S41 | MA telemedicine | 36 | e-counseling.sh. |
|  | #34 | MeSH descriptor:  [Internet-Based Intervention]  explode all trees | 36 | Internet-Based Intervention  [MeSH Terms] | S42 | MA internet-based intervention | 37 | “intervention”.ti. or “intervention”.ab. |
|  | #35 | MeSH descriptor: [Distance Counseling] explode all trees | 37 | Distance counseling  [MeSH Terms] | S43 | MA distance counseling | 38 | “treatment”.ti. or “treatment”.ab. |
|  | #36 | (“intervention”):ti,ab,kw | 38 | “intervention” [Title/Abstract] | S44 | SU treatment | 39 | “program”.ti. or “program”.ab. |
|  | #37 | (“treatment”):ti,ab,kw | 39 | “treatment” [Title/Abstract] | S45 | SU psychotherapy | 40 | “therap*”.ti. or “therap*”.ab. |
|  | #38 | (“program”):ti,ab,kw | 40 | “program” [Title/Abstract] | S46 | SU couples therapy | 41 | “psychotherap*”.ti. or “psychotherap*”.ab. |
|  | #39 | (therap*):ti,ab,kw | 41 | “therap*” [Title/Abstract] | S47 | SU counseling | 42 | “couple therap*”.ti. or  “couple therap*”.ab. |
|  | #40 | (psychotherap*):ti,ab,kw | 42 | “psychotherap*” [Title/Abstract] | S48 | SU psychosocial readjustment | 43 | “counsel*”.ti. or “counsel*”.ab. |
|  | #41 | (counsel*):ti,ab,kw | 43 | “counsel*” [Title/Abstract] | S49 | SU computer Assisted Therapy | 44 | “support service*”.ti. or  “support service*”.ab. |
|  | #42 | (support service*):ti,ab,kw | 44 | “support service*” [Title/Abstract] | S50 | SU telemedicine | 45 | “advocacy”.ti. or “advocacy”.ab. |
|  | #43 | (“advocacy”):ti,ab,kw | 45 | “advocacy” [Title/Abstract] | S51 | SU electronic health services | 46 | “secondary prevention”.ti. or “secondary prevention”.ab. |
|  | #44 | (“secondary prevention”):ti,ab,kw | 46 | “secondary prevention” [Title/Abstract] | S52 | SU digital interventions | 47 | “tertiary prevention”.ti. or  “tertiary prevention”.ab. |
|  | #45 | (“tertiary prevention”):ti,ab,kw | 47 | “tertiary prevention” [Title/Abstract] | S53 | SU mobile health | 48 | “rehabilitation”.ti. or “rehabilitation”.ab. |
|  | #46 | (“rehabilitation”):ti,ab,kw | 48 | “rehabilitation” [Title/Abstract] | S54 | TI “intervention” OR  AB “intervention” |  |  |
|  |  |  |  |  | S55 | TI “treatment” OR  AB “treatment” |  |  |
|  |  |  |  |  | S56 | TI “program” OR AB “program” |  |  |
|  |  |  |  |  | S57 | TI “therap*” OR AB “therap*” |  |  |
|  |  |  |  |  | S58 | TI „psychotherap*“ OR  AB “psychotherap*“ |  |  |
|  |  |  |  |  | S59 | TI “cousel*” OR AB “cousel*” |  |  |
|  |  |  |  |  | S60 | TI “support service*” OR  AB “support service*” |  |  |
|  |  |  |  |  | S61 | TI “advocacy” OR  AB “advocacy” |  |  |
|  |  |  |  |  | S62 | TI “empowerment” OR  AB “empowerment” |  |  |
|  |  |  |  |  | S63 | TI “secondary prevention” OR AB “secondary prevention” |  |  |
|  |  |  |  |  | S64 | TI “tertiary prevention” OR  AB “tertiary prevention” |  |  |
|  |  |  |  |  | S65 | TI “rehabilitation” OR  AB “rehabilitation” |  |  |
|  | **#47** | #25 OR #26 OR #27 OR #28 OR #29 OR #30 OR #31 OR #32 OR #33 OR #34 OR #35 OR #36 OR #37 OR #38 OR #39 OR #40 OR #41 OR #42 OR #43 OR #44 OR #45 OR #46 | **49** | 27 OR 28 OR 29 OR 30 OR 31 OR 32 OR 33 OR 34 OR 35 OR 36 OR 37 OR 38 OR 39 OR 40 OR 41 OR 42 OR 43 OR 44 OR 45 OR 46 OR 47 OR 48 | **S66** | S33 OR S34 OR S35 OR S36 OR S37 OR S38 OR S39 OR S40 OR S41 OR S42 OR S43 OR S44 OR S45 OR S46 OR S47 OR S48 OR S49 OR S50 OR S51 OR S52 OR S53 OR S54 OR S55 OR S56 OR S57 OR S58 OR S59 OR S60 OR S61 OR S62 OR S63 OR S64 OR S65 | **49** | 27 OR 28 OR 29 OR 30 OR 31 OR 32 OR 33 OR 34 OR 35 OR 36 OR 37 OR 38 OR 39 OR 40 OR 41 OR 42 OR 43 OR 44 OR 45 OR 46 OR 47 OR 48 |
|  | #48 | MeSH descriptor:  [Randomized Controlled Trials as Topic] explode all trees | 50 | randomized controlled trials as topic [MeSH Terms] | S67 | MA randomized controlled trials | 50 | randomized controlled trial.sh. |
|  | #49 | MeSH descriptor:  [Clinical Trials as Topic]  explode all trees | 51 | clinical trials as topic  [MeSH Terms] | S68 | MA clinical trials | 51 | clinical trial.sh. |
|  | #50 | (“randomized controlled trial”):pt | 52 | “randomized controlled trial” [Publication Type] | S69 | SU randomized clinical trial | 52 | RCT.ti. or RCT.ab. |
|  | #51 | (“controlled clinical trial”):pt | 53 | “clinical trial” [Publication Type] | S70 | SU treatment effectiveness evaluation | 53 | random*.ti. or random*.ab. |
|  | #52 | (clinical trial):pt | 54 | “controlled clinical trial” [Publication Type] | S71 | TI RCT OR AB RCT | 54 | trial.ti. or trial.ab. |
|  | #53 | (clinical trial protocol):pt | 55 | “clinical trial” [Publication Type] | S72 | TI random* OR AB random* | 55 | control condition.ab. |
|  | #54 | (RCT):ti,ab,kw | 56 | “clinical trial protocol”  [Publication Type] | S73 | TI trial OR AB trial |  |  |
|  | #55 | (clinical trial):ti,ab,kw | 57 | “clinical study” [Publication Type] | S74 | TI effectiveness OR  AB effectiveness |  |  |
|  | #56 | (random*):ti,ab,kw | 58 | RCT [Title/Abstract] | S75 | TI experiment* OR  AB experiment* |  |  |
|  | #57 | (“trial”):ti, ab, kw | 59 | random* [Title/Abstract] |  |  |  |  |
|  | #58 | (“study”):ti,ab,kw | 60 | trial [Title/Abstract] |  |  |  |  |
|  | **#59** | #48 OR #49 OR #50 OR #51 OR #52 OR #53 OR #54 OR #55 OR #56 OR #57 OR #58 | **61** | 50 OR 51 OR 52 OR 53 OR 54 OR 55 OR 56 OR 57 OR 58 OR 59 OR 60 | **S76** | S67 OR S68 OR S69 OR S70 OR S71 OR S72 OR S73 OR S74 OR S75 | **56** | 48 OR 49 OR 50 OR 51 OR 52 OR 53 OR 54 OR 55 |
|  | **#60** | **#24 AND #47 AND #59** | **62** | **26 AND 49 AND 61** | **S77** | **S32 AND S66 AND S76** | **58** | **27 AND 49 AND 56** |

### Appendix B

| **Domain** | **Items** |
| --- | --- |
| Study identification characteristics | - Study identifier (first author, year) - Author(s) - Publication year - Title - Type of record - Corresponding records |
| Study design  characteristics | - Trial location - Recruiting strategy - Trial setting - Total number of groups and times of measurements - Timing of measurements (period from baseline in weeks) - Primary and secondary outcomes - Randomization method (including unit of allocation) - Sample size (total and per group) - Dropout (per group and timing of measurement) - Numbers analyzed (per group and timing of measurement) - Whether reasons for dropout were reported - Type of analysis (i.e., per-protocol and/or intention-to-treat analysis including imputation strategy) |
| Participant  characteristics | - Age (overall and per group) - Type of gender included (including proportion of genders included per group) - Type of target population - Baseline experience of IPV (including duration of exposure to IPV) - Significant baseline differences between study groups - Inclusion and exclusion criteria |
| Intervention characteristics | - Number of intervention groups - Name and description of intervention(s) - Theoretical approach (for each intervention) - Delivery mode (for each intervention) - Facilitator(s) of intervention(s) - Number, duration, and frequency of intervention sessions - Total intervention duration |
| Control  characteristics | - Number of control groups - Type of control group(s) (e.g., waitlist, TAU, placebo, active control) - Description of control condition(s) |
| Outcomes | - Definition of outcome - Outcome measure(s) used (including whether low or high scores are good) - Timing of measurements - Raw data (for each group and each time point, e.g., means, SD, SE, t-value)   If applicable for each outcome of the prespecified domains: safety-related outcomes (IPV, safety behavior), mental health outcomes (depression, anxiety, PTSD, substance use), psychosocial outcomes (self-efficacy, self-esteem, social support, decisional conflict, empowerment, quality of life), service use, satisfaction, acceptance, other relevant outcomes (e.g., suicidality, psychological distress, general mental health), treatment, adherence, adverse events |
| Conclusions | Key conclusions of author(s) |
